# GWAS Meta-analysis Identifies Novel Associated Loci and Points to Causal Tissues in Central Serous Chorioretinopathy

**DOI:** 10.64898/2026.05.20.26353693

**Authors:** Liyin Chen, Soo Hyun Kim, Buu Truong, Joel T Rämö, Bryan R Gorman, Elon H. C. van Dijk, Joost Brinks, Tiit Nikopensius, Seung Hoan Choi, Risto Kajanne, Juha Mehtonen, Kai Kaarniranta, Lucia Sobrin, Mitja Kurki, Suzanne Yzer, VA Million Veteran Program, FinnGen, Wen-Chih Wu, Joni A Turunen, Ayellet J Segrè, Josep Maria Mercader, Alicia Huerta, Mark J Daly, Aarno Palotie, Patrick T Ellinor, Camiel JF Boon, Sudha K Iyengar, Neal S Peachey, Pradeep Natarajan, Elizabeth J Rossin

## Abstract

**Objective:** To define CSC genetic architecture and identify implicated ocular tissues, cell types, genes, and circulating proteins.

**Data Sources:** Genome-wide data were assembled from FinnGen, All of Us, Mass General Brigham Biobank, Million Veteran Program, and a Dutch chronic CSC cohort. Serum protein quantitative trait loci, human single-cell ocular atlases, and UK Biobank macular optical coherence tomography (OCT) imaging were used for downstream analyses.

**Study Selection:** Five European-ancestry cohorts with genome-wide data and cohort-specific CSC case-control definitions were included, comprising 2,584 cases and 1,044,455 controls. Variants present in at least 2 cohorts were meta-analyzed.

**Data Extraction and Synthesis:** Cohort-level GWASs were adjusted for age, age squared, sex, genotyping array or batch, and 10 genetic principal components, then combined using fixed-effects inverse-variance meta-analysis. Post-GWAS analyses included gene prioritization, colocalization, Mendelian randomization, single-cell disease-relevance scoring, and testing of a CSC genetic risk score in UK Biobank OCT images.

**Main Outcome(s) and Measure(s):** Genome-wide significant CSC loci, effector genes and proteins, tissue and cell-type enrichment, and CSC-relevant OCT abnormalities.

**Results:** Across 11,068,938 variants, 10 loci reached genome-wide significance (*P* < 5 × 10−8), including 3 novel loci near *TGFB1, LINC00551*, and *LOC105375630* and 7 replicated loci near *CFH, CD46, NOTCH4, PREX1, PTPRB, GATA5*, and *TNFRSF10A*. Integrative analyses prioritized 10 candidate effector genes. Colocalization and Mendelian randomization implicated circulating TNFRSF10A, TGFB1, and CASP10 levels. Single-cell analyses localized genetic risk to sclera (*P* = 2.0 × 10−4) and vascular endothelial cells (*P* = 4.0 × 10−4), with fibroblast enrichment. In UK Biobank, OCT abnormalities were more frequent in the top vs bottom 1% of CSC genetic risk (18 of 109 [16.5%] vs 8 of 134 [6.0%]; odds ratio, 4.05; 95% CI, 1.65-10.87; *P* = .002).

**Conclusions and Relevance:** In this GWAS meta-analysis, CSC susceptibility localized predominantly to scleral and vascular biology rather than primary retinal pigment epithelial dysfunction. These findings support CSC as a sclerovascular disorder and nominate complement regulation, endothelial signaling, and extracellular matrix pathways for future study.

**Key Points:** *Question:* What genetic loci, genes, proteins, and ocular cell types underlie susceptibility to central serous chorioretinopathy (CSC)?

*Findings:* In this GWAS meta-analysis of 2,584 CSC cases and 1,044,455 controls, 10 genome-wide significant loci were identified, including 3 novel loci. Integrative analyses implicated scleral fibroblasts and vascular endothelial cells, prioritized candidate effector genes and circulating proteins, and showed that high CSC genetic risk was associated with more frequent RPE abnormalities on macular OCT.

*Meaning:* These findings support CSC as a primary sclerovascular disorder and nominate mechanism-linked pathways for future translational studies.

*Importance:* The primary site of dysfunction in central serous chorioretinopathy (CSC) remains uncertain.

## Introduction

Central serous chorioretinopathy (CSC) is an important cause of central vision loss in workin-gaged adults, but its pathophysiology remains incompletely understood. CSC is characterized by accumulation of subretinal fluid, detachments of the retinal pigment epithelium (RPE), and choroidal thickening (Feenstra et al., 2024). Symptoms can involve decreased visual acuity, central scotoma, micropsia, or metamorphopsia (Wang *et al*., 2008). Disease onset is usually between 20 and 50 years of age with males more commonly affected by up to 6-fold (Kitzmann *et al*., 2008; Fung, Yang and Kam, 2023). CSC remains a multifactorial disease with associated risk factors including corticosteroid use, type A behavior, obstructive sleep apnea, *Helicobacter pylori* infection, uncontrolled hypertension, and pregnancy, along with other vascular disorders. At the anatomic level, CSC is associated with abnormally dilated and leaking choroidal vasculature (Daruich et al., 2015; Brinks et al., 2022; Pauleikhoff et al., 2024), RPE dyfunction and disruption, and abnormally thickened sclera (Imanaga et al., 2021).

Recent advances in population genetics have offered novel insights into the etiology of CSC. Previous studies found that about half of CSC patients had at least one relative also present with CSC symptoms, suggesting a genetic predisposition (Weenink, Borsje and Oosterhuis, 2001; van Dijk *et al*., 2019). Genome-wide association studies (GWAS) in European (EUR) (Schellevis *et al*., 2018a) and Japanese (Miki *et al*., 2014; Hosoda *et al*., 2019) patients have identified loci near *complement factor H (CFH*), *GATA5*, and *TNFRSF10A*. We have previously published a 3-cohort meta-analysis of CSC that identified novel loci near *PREX1, CD46*, and *NOTCH4*, pointing to the likely role of endothelial dysfunction (Rämö *et al*., 2023). Following this, a large meta-analysis showed a strong association with the specific missense variant rs113791087 in *PTPRB* that co-associates with varicose veins, underscoring the likely vascular nature of the disease (Rämö, Gorman, *et al*., 2025a). A recent meta-analysis of Japanese and EUR cohorts confirmed involvement of *CFH, TNFRSF10A*, and *GATA5* but was unable to replicate other loci (Mori *et al*., 2025). Overall, the current literature remains underpowered due to modest case counts. Moreover, most studies have limited post-GWAS analyses to nominate causal genes and cell types, limiting the utility of these studies.

To address these limitations, we performed a EUR CSC case-control GWAS meta-analysis with an integrated post-GWAS framework tailored to CSC biology. We aggregated five cohorts, specifically FinnGen (Kurki *et al*., 2023), All of Us (AoU) research program (All of Us Research Program Investigators *et al*., 2019), Mass General Brigham Biobank (MGBB) (Castro *et al*., 2022; Koyama *et al*., 2025), Million Veteran Program (MVP) (Gaziano *et al*., 2016), and a previously published Dutch cohort (Schellevis *et al*., 2018b). We then deployed complementary post-GWAS analyses including two-sample Mendelian randomization (MR) and Bayesian colocalization with cis-pQTL resources to test whether genetically proxied circulating protein changes are causal for CSC. Single-cell disease-relevance scoring across ocular atlases then localized genetic risk to discrete ocular cell types and tissues, and a genetic-risk-to-ocular-anatomy bridge in UK Biobank (UKB) optical coherence tomography (OCT) evaluated whether polygenic burden manifests as measurable alterations on macular OCT.

## Methods

### Study design

We conducted a GWAS meta-analysis of CSC using five European-ancestry cohorts: FinnGen, AoU, MGBB, MVP, and a previously reported Dutch chronic CSC cohort. FinnGen is a public-private partnership research project that combines genotype data from newly collected and legacy samples administered by Finnish biobanks (https://www.finngen.fi/en). This study included genotype data from 486,484 individuals from FinnGen Data Freeze 12. The MGBB is a biorepository patient samples at Mass General Brigham (Koyama *et al*., 2025). We included 53,846 genotyped participants in MGBB. In AoU, we used genetic data from 228,707 short-read whole genome-sequencing samples in the v8 release and excluded participants without linked electronic health record data. The Dutch chronic-CSC (cCSC) cohort included 521 cases and 3,577 controls. More details for each cohort are provided in Supplementary Information.

UKB is a deeply phenotyped and genotyped prospective population-level cohort (Sudlow *et al*., 2015) for which a subset of participants has OCT. This research has been conducted using the UK Biobank Resource under Application Number 50211.

### Ethics statement

The MGBB study protocol was approved by the MGB Institutional Review Board. Use of All of Us data for this study was approved by the All of Us research program after completing all required training. The study of European patients with cCSC was carried out in accordance with the tenets of the Declaration of Helsinki and was approved by the local ethics committees of the Radboud University Medical Center, Leiden University Medical Center, and University Hospital of Cologne. Written informed consent was obtained for all MGBB, All of Us, Dutch cCSC participants. The MVP024 study protocol was approved by the Veterans Affairs central Institutional Review Board. Complete ethics statement of FinnGen is provided in Supplementary Information.

### Phenotype ascertainment

CSC was defined using cohort-specific electronic health record codes or previously published phenotype definitions. In MGBB, CSC diagnoses identified by ICD-9-CM or ICD-10-CM codes were validated by retina-specialist review of OCT images, with fluorescein or indocyanine green angiography reviewed when available. Participants with age-related macular degeneration were excluded from CSC cases and controls. Detailed case-control definitions are provided in Supplementary Information.

### Genome-wide Association Studies and Meta-analysis

The GWAS in FinnGen, AoU, MVP and MGBB were performed using REGENIE (v3, v2.2.4 for FinnGen) (Mbatchou *et al*., 2021), limiting to European samples (ancestry definitions for each cohort are described in Supplementary Information). The GWAS for the Dutch cohort was described previously (Schellevis *et al*., 2018b). All analyses were adjusted for age, age-squared, sex, genotyping array/batch, and the first 10 genomic PCs. Genomic control inflation (λ_GC_) was computed to assess systemic confounding in each cohort.

We combined cohort-level summary statistics using the Genome Wide Association Meta Analysis tool GWAMA (v2.2.2) (Mägi and Morris, 2010). Variants were included if present in at least 2 cohorts. 11,068,938 rsID-labeled SNPs were used for subsequent analyses. Locus definition followed a ±500 kb window around each lead SNP with LD r^2^ ≥ 0.05 using 1000 Genomes Phase 3 (1000 Genomes Project Consortium *et al*., 2015) European (EUR) samples as LD reference.

### Post-GWAS analyses

The details of MAGMA, Polygenic Priority Score (PoPS), colocalization analysis, mendelian randomization and single-cell disease relevance score analysis (scDRS) can be found in the supplement and followed standard default protocols.

### Genetic risk score (GRS) and OCT in UKB

We constructed a CSC GRS using the 10 genome-wide significant lead variants identified in the CSC meta-analysis (Table 1), weighted by meta-analysis log-odds ratios. We matched effect alleles between the meta-analysis and UKB genotypes, removed strand-ambiguous markers without clear MAF alignment and computed per-individual GRS using the *--score* function in PLINK 1.9 (Igo, Kinzy and Cooke Bailey, 2019). We restricted the analysis to unrelated European-ancestry participants with array genotype data and macular OCT scans. Participants in the top and bottom 1% of the GRS distribution were selected for imaging review by a retinal specialist.

**Table 1:** Ten genome-wide significant loci in CSC GWAS meta-analysis. Results are derived from a fixed-effects inverse-variance-weighted meta-analysis of five European-ancestry cohorts. Each cohort-level GWAS was performed with adjustment for age, age^2^, sex, genotyping array/batch, and the first 10 genomic principal components. Variants meeting genome-wide significance (P < 5 × 10^−^ □) and present in ≥2 cohorts are shown; novel loci not previously reported in CSC are denoted with an asterisk (*). Abbreviations: rsID, reference SNP cluster identifier; CHR, chromosome; position, genomic position in GRCh38 coordinates (base pairs); A1, effect allele; beta, per-allele log-odds ratio; se, standard error; p-value, fixed-effects meta-analysis P value; Heterogeneity P, P value for Cochran’s Q test of between-study heterogeneity.

| rsID | Nearest gene | CHR | position | A1 | beta | se | p-value | Heterogeneity P |
| --- | --- | --- | --- | --- | --- | --- | --- | --- |
| rs10922105 | CFH | 1 | 196721120 | C | 0.349 | 0.0281 | 3.31E-35 | 0.102 |
| rs2724360 | CD46 | 1 | 207769813 | T | 0.297 | 0.0367 | 6.46E-16 | 0.558 |
| rs113791087 | PTPRB | 12 | 70559589 | G | 1.109 | 0.141 | 4.82E-15 | 0.279 |
| rs8192569 | NOTCH4 | 6 | 32222707 | A | 0.319 | 0.0467 | 9.09E-12 | 0.396 |
| rs2427357 | GATA5 | 20 | 62450103 | C | 0.221 | 0.0325 | 1.20E-11 | 0.255 |
| rs73045269* | TGFB1, CCDC97 | 19 | 41319286 | T | -0.264 | 0.0402 | 5.02E-11 | 0.547 |
| rs34822542 | PREX1 | 20 | 48641217 | C | 0.2407 | 0.0401 | 2.04E-09 | 0.280 |
| rs13278062 | TNFRSF10A | 8 | 23225458 | T | 0.165 | 0.0286 | 9.17E-09 | 0.312 |
| rs150013629* | LOC105375630 | 8 | 88447463 | G | 1.197 | 0.2182 | 4.17E-08 | 0.228 |
| rs1888541* | LINC00551 | 13 | 106697763 | A | 0.219 | 0.0400 | 4.53E-08 | 0.888 |

Macular spectral-domain OCT scans (Topcon 3D OCT-1000) from the selected high- and low-risk participants underwent manual grading by an attending retina specialist. The grader (EJR) and analyst (LC) were masked to the participants’ GRS status to prevent bias. The primary outcome was the presence of any RPE abnormalities seen in CSC in either eye including RPE detachment(s) or other RPE irregularities, evidence of pattern dystrophy or pattern-like changes, subretinal fluid, evidence of CSC, pachychoroid pigment epitheliopathy or outer retinal atrophy. Unrelated findings like epiretinal membrane, drusen, or other subtle abnormalities were not counted. Scans were binarized as “abnormal” or “normal” based on the presence of one or more of the aforesaid primary outcome findings. Differences in the proportion of participants with OCT abnormalities between the top and bottom 1% GRS percentiles were assessed by multivariable logistic regression adjusted for age, sex, and the first 10 genomic principal components, with the likelihood-ratio test as the primary inference. Statistical significance was defined as a 2-sided P < 0.05.

### Effect size comparison between CSC and AMD

Because the CSC GWAS is still relatively underpowered for formal genetic correlation analysis, we compared effect sizes at CSC loci identified here and previously reported AMD loci (Fritsche *et al*., 2016). For each locus, the lead variant was used when available in both datasets; otherwise, a proxy variant in linkage disequilibrium was chosen. Proxy variants were precomputed using the 1000 Genomes Phase 3 EUR reference panel within ±50 kb windows around each lead variant and ranked by linkage disequilibrium (R^2^), with the highest-R^2^ proxy selected among those present in both datasets. Only loci for which either the lead variants or eligible proxies were available and achieved nominal significance (P < 0.05) in both datasets were included. Effect sizes were harmonized to a common effect allele prior to analysis. Pearson correlation coefficients and linear regression lines were computed to quantify cross-trait concordance.

## Results

### Meta-analysis and cohort-level GWAS

Across the 5 cohorts, we analyzed 2,584 CSC cases and 1,044,455 controls from European ancestry (Supplementary Data 1 and 2). 11,068,938 variants were included for meta-analysis after quality control. Fixed-effects meta-analysis yielded 10 loci reaching genome-wide significance (*P* < 5 x 10^8^) (Figure 1, Table 1). Seven loci confirmed previous associations: variants near *CFH, CD46, NOTCH4, PREX1, PTPRB* and *GATA5* had been identified by our group before. Notably, we replicated a variant near *TNFRSF10A* for the first time in a European cohort, validating a risk locus that was originally identified only in Japanese cohorts. Three novel genome-wide significant loci were also identified, with nearest genes *TGFB1, LINC00551*, and *LOC105375630*. Effect sizes and directions were similar for each locus in each cohort (Figure 1b, P-value for Cochran’s Q test statistic for heterogeneity > 0.1). The quantile–quantile plot showed modest deviation from the null consistent with polygenicity (λ_GC_ = 1.055, Supplementary Figure S1 and S2).

**Figure 1.**
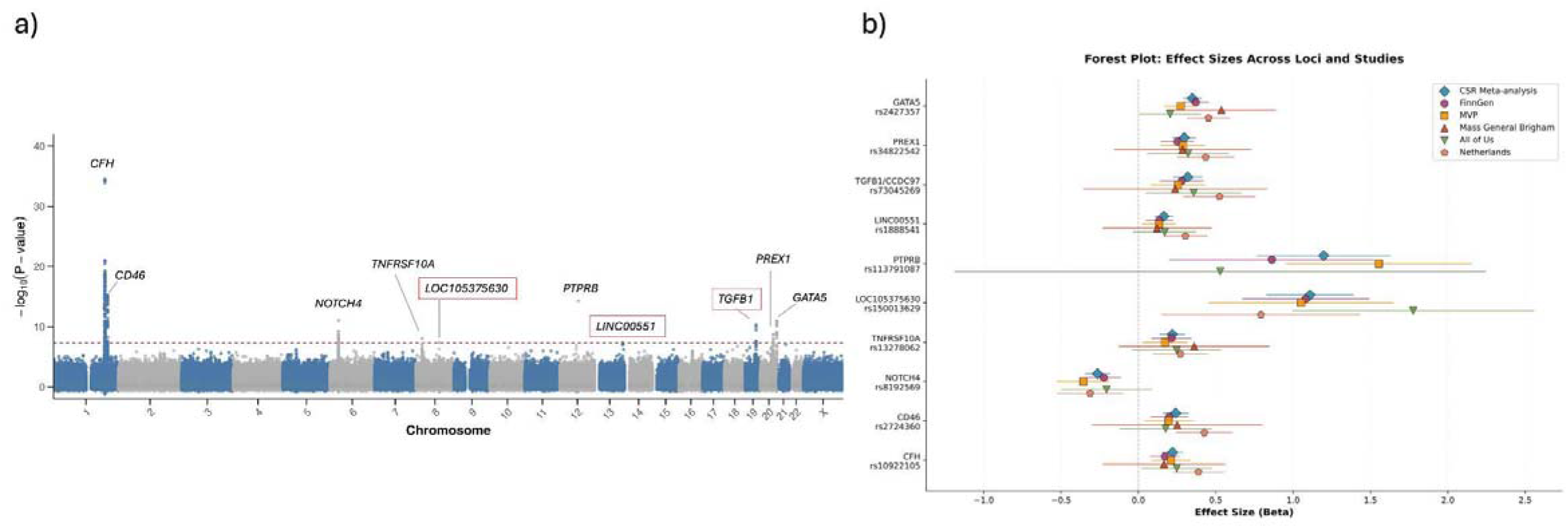
Genome-wide association meta-analysis of central serous chorioretinopathy identifies 10 associated loci. (a) Manhattan plot of −log_10_(P) and chromosomal position for all variants present in the meta-analysis. Each point represents a single variant; chromosomes alternate in color. The horizontal red line indicates the genome-wide significance threshold (P = 5 × 10^−^□). The nearest gene is annotated for each genome-wide significant locus; novel loci are labeled in red boxes. (b) Forest plots of per-allele effect sizes (log-odds ratios) in individual GWASs and the meta-analysis. Horizontal lines indicate 95% confidence intervals.

### Gene prioritization

We performed gene-based association analysis using MAGMA. 19 genes passed the Bonferroni-corrected threshold (P < 0.05/17,832), among which 4 of them (*CD46, GATA5, PREX1*, and *CFH*) were nearest to the identified lead genome-wide significant SNP (Supplementary Data 3).

We applied the PoPS framework to propose effector genes with biological relevance. Of the 10 GWAS loci, all but 1 locus had at least 1 positively scored gene from PoPS (Supplementary Data 4). PoPS prioritized genes agreed with the nearest-gene assignment for 7 of the 10 loci, including *CFH, TNFRSF10A*, and *TGFB1*.

### Colocalization and MR with serum protein levels

To test whether any CSC variants modulate CSC risk through serum protein levels, we first performed Bayesian colocalization with 2,923 serum proteins using the pQTL dataset from the UKB (Sun *et al*., 2023) and our CSC GWAS meta-analysis. We then carried out two-sample MR restricted to the proteins with strong colocalization evidence to test the direction and magnitude of the genetically proxied effect.

We identified strong evidence of colocalization (PP4 > 0.90) between the CSC GWAS signal and the pQTL signals for 3 proteins, namely TNFRSF10A, TGFB1, and CASP10 (PP4>0.95 for all three, Supplementary Data 5). LocusCompare visualizations confirmed that the same high-LD variant clusters drove both the pQTL and CSC GWAS signals for these proteins, verifying that the colocalization results reflect shared genetic architecture (Figure 2a).

**Figure 2.**
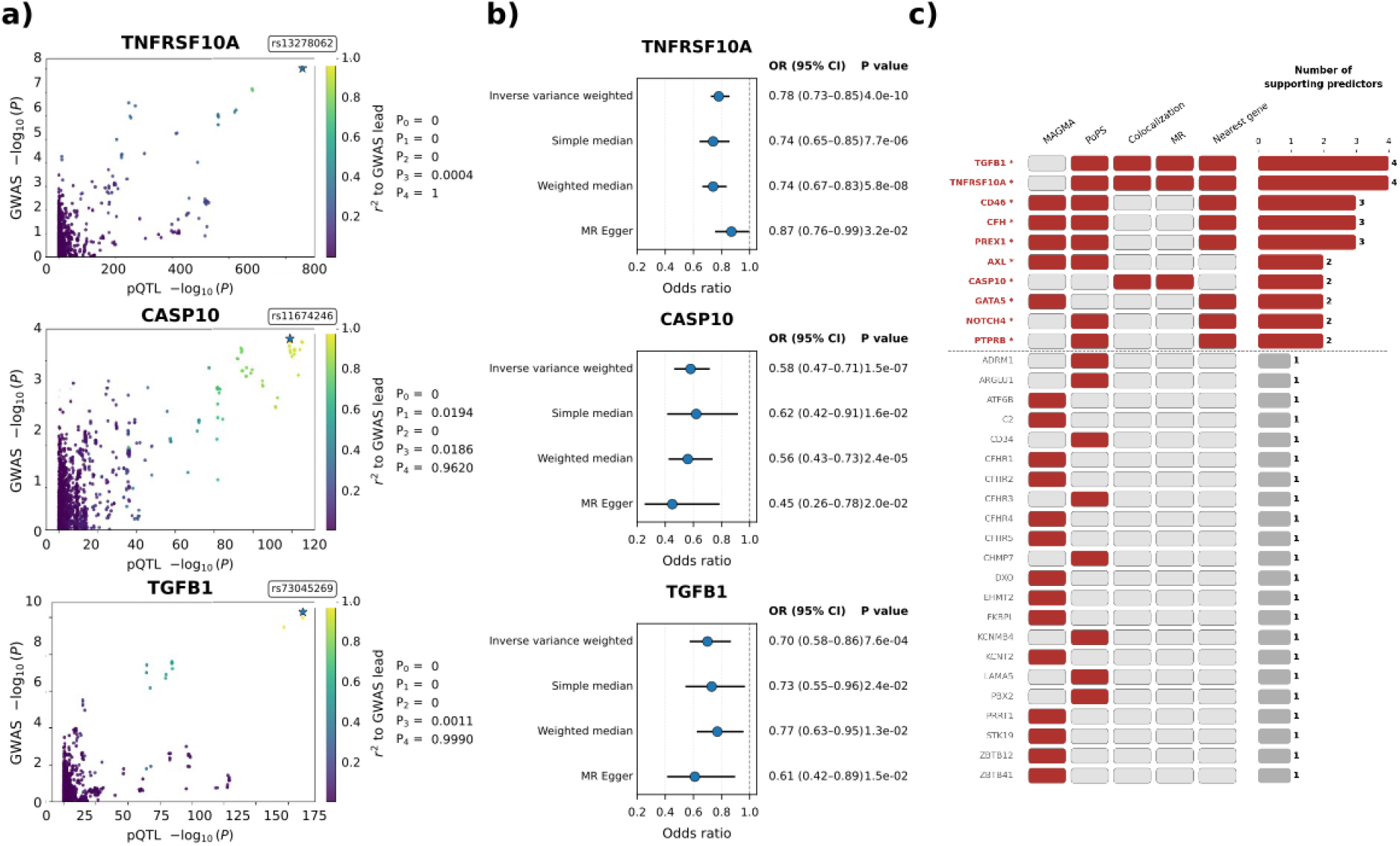
Convergent genetic, proteomic, and causal evidence prioritizes candidate genes for CSC. (a) LocusCompare plots of the 3 proteins with significant colocalization (PP4 > 0.95) between the cis-pQTL and the CSC GWAS signal. x-axis shows the P value of each variant in the serum protein pQTL, and the y-axis shows the P value of each variant in the CSC meta-analysis. The right column reports the posterior probability estimates from coloc. b) Forest plot of MR estimates for the same proteins shown in (a). For each protein, points show the causal odds ratio (OR) for CSC per genetically predicted increase in protein level. Rows within each panel correspond to complementary MR estimators. The vertical dashed line marks the null (OR = 1). The two right columns report the OR (95% CI) and the P value for each method (Bonferroni-corrected significance threshold: P < 0.05/3 = 0.0167). c) Binary heatmap showing the mapping of candidate genes (rows) to 5 gene prioritization criteria (columns): MAGMA, PoPS, colocalization, MR, and nearest gene. Red cells indicate that a gene was identified by the corresponding method; grey cells indicate no support. Genes supported by 2 or more independent predictors (above dashed line, bold red labels with asterisks) were designated as prioritized genes. Of 33 candidate genes, ten were prioritized based on convergent evidence.

Because colocalization does not necessarily indicate causality, we performed MR to test whether altered serum protein levels are in the causal pathway to CSC for the 3 proteins related to the association signals that colocalized. Among the 3, MR estimates were directionally consistent across all complementary estimators. Genetically predicted higher circulating TNFRSF10A was associated with reduced CSC risk and passed the Bonferroni-corrected threshold of P < 0.05/3 = 0.0167 (Figure 2b). TGFB1 and CASP10 were also significant at this threshold, with genetically predicted higher levels of each associated with lower CSC risk (Figure 2b).

Effect-size estimates for all 3 proteins were concordant in direction and similar in magnitude across complementary MR estimators (inverse-variance–weighted, simple median, and weighted median, MR-Egger), with no evidence of significant directional horizontal pleiotropy (MR-Egger intercept P > 0.05) (Figure 2b) (Supplementary Data 6).

### Endothelial and scleral enrichment in scRNA-seq data

To elucidate the cellular basis of CSC genetic risk, we applied scDRS to 3 complementary human single-cell atlases using a combined set of genes prioritized from the genetic analyses discussed above. The results revealed a specific enrichment in scleral and vascular tissues (Supplementary Data 7).

At the tissue level, sclera was enriched (Figure 3). In the posterior segment atlas, scleral tissue achieved the highest score of all tested tissues (P = 2.0 × 10^-4^; z = 4.87; FDR < 0.05). This finding was replicated in the multi-organ Tabula Sapiens atlas, where the sclera again was enriched compared to all other ocular tissues (P = 0.0036; z = 3.70; FDR < 0.05). Additionally, the scleral lamina cribrosa, the scleral structure of the optic nerve head, demonstrated nominal significant enrichment (P = 0.037; z = 2.00; FDR < 0.1). Of note, vascular tissue is not a separate tissue category because it is not separated mechanically from other tissues.

**Figure 3.**
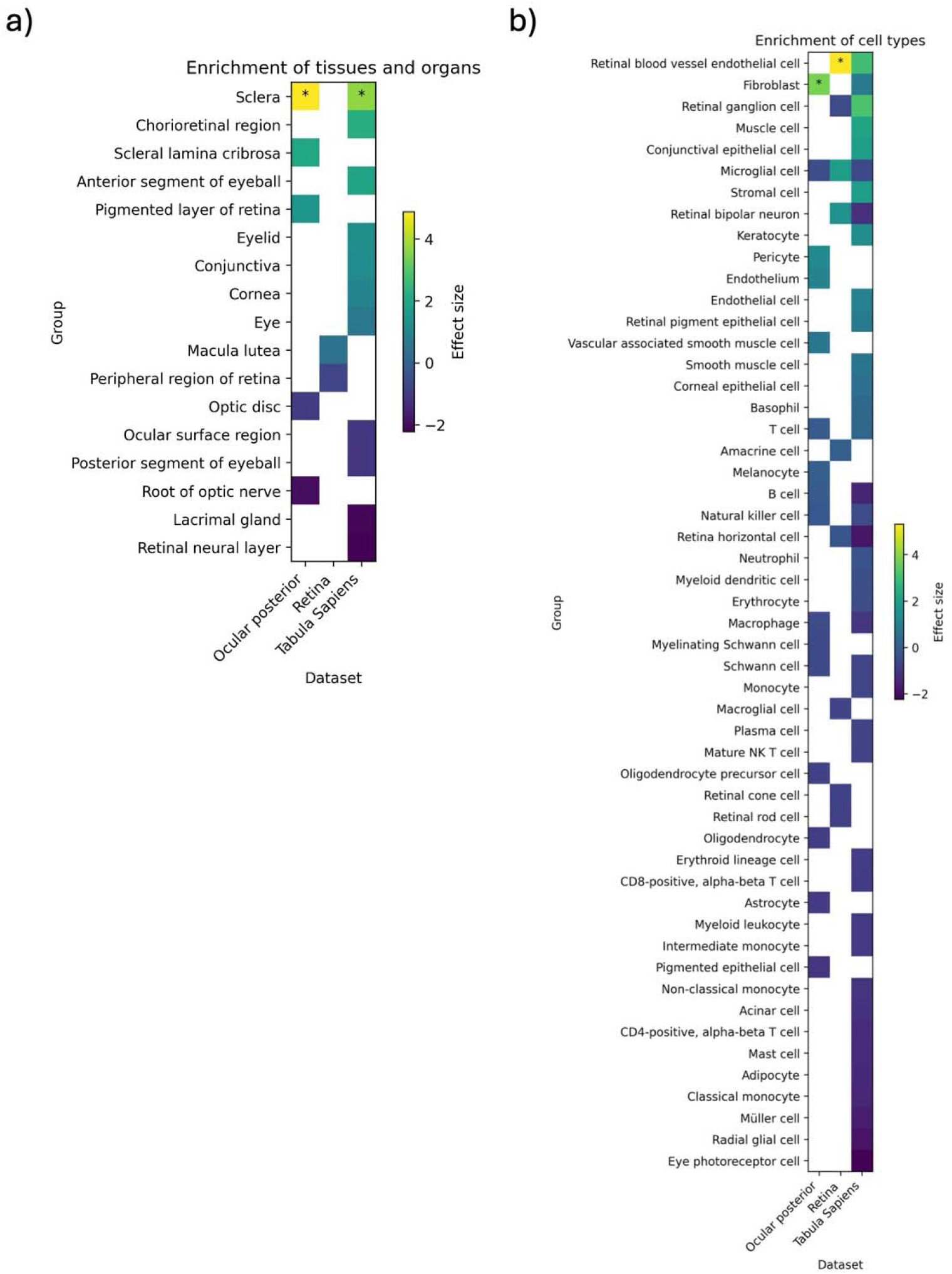
Single-cell disease relevance scoring reveals vascular and scleral enrichment of central serous chorioretinopathy (CSC) genetic risk. Heatmap displaying Monte Carlo z-scores from scDRS analysis of CSC GWAS summary statistics across (a) anatomical regions and tissue types and (b) cell types. Columns represent 3 independent datasets: Monavarfeshani et al. (posterior segment atlas), Menon et al. (retinal atlas), and Tabula Sapiens (multi-organ atlas). Rows indicate anatomical groupings and tissue types. Color intensity represents the strength and direction of genetic association, with warm colors (yellow-green) indicating positive enrichment of CSC genetic risk and cool colors (purple-blue) indicating depletion. Asterisks denote FDR < 0.05.

At the single cell level (whereby tissues are dissociated and homogenized into single cells), vascular endothelial cells and fibroblasts were identified as significant cellular expressors of CSC risk genes among all cell types in the two single-cell datasets containing these cells. In the specialized retinal atlas, retinal blood vessel endothelial cells were enriched (P = 4.0 × 10^-4^; z=5.31; FDR < 0.05). This was corroborated by the Tabula Sapiens atlas, where retinal blood vessel endothelial cells (P = 0.0058; z = 3.01; FDR = 0.11) exhibited the most substantial enrichment signal among the 52 cell types in the dataset. Fibroblasts, a key structural component of the sclera, displayed significant enrichment in the posterior segment atlas (P = 4.0 × 10^-4^; z = 3.81; FDR < 0.05) (Supplementary Figure S3).

In contrast to the robust vascular and scleral signals, cell types intrinsic to the neurosensory retina, immune system and RPE showed no significant evidence of expressing genes associated to CSC. Furthermore, immune populations, including T cells, B cells, macrophages, and neutrophils, consistently yielded null or negative association scores across all datasets (P > .05 for all immune subsets).

### CSC GRS associated with RPE abnormality in UKB

To evaluate whether polygenic burden for CSC manifests as measurable anatomic changes in an unselected population, we constructed a CSC GRS from genome-wide significant independent variants and applied it to unrelated European-ancestry UKB participants with macular OCT imaging. We evaluated macular OCT scans from 243 participants drawn from a masked pool comprising the top 1% (high risk; n = 109) and bottom 1% (low risk; n = 134) of the CSC GRS distribution. The proportion of participants with at least one RPE abnormality in either eye (see Methods) was approximately 2.6-fold higher in the high-risk group than in the low-risk group (16.51% [18/109] vs 5.97% [8/134]). In multivariable logistic regression adjusted for age, sex, and the first 10 genomic principal components, high GRS status was associated with a nearly four-fold increase in the odds of an abnormal OCT (OR = 4.05, 95% CI 1.65-10.87; likelihood-ratio P = 2.0 × 10^−3^). Representative high-GRS scans are shown in Figure 4.

**Figure 4.**
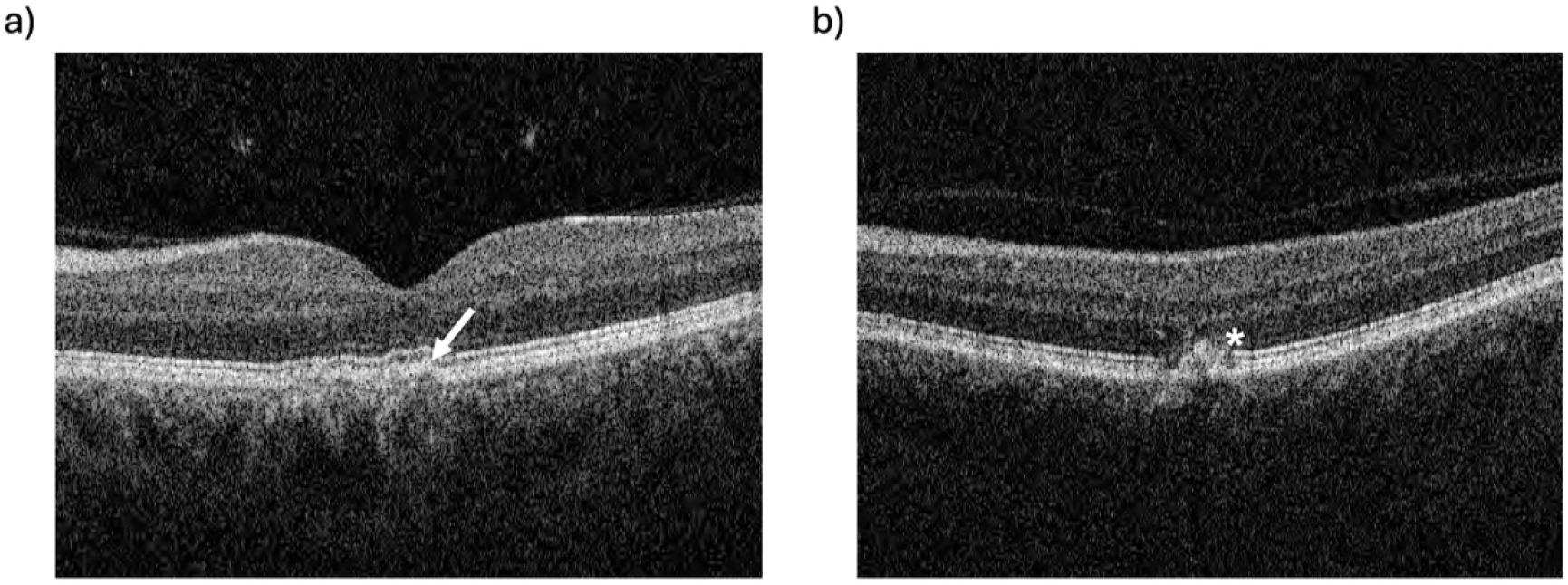
Representative abnormal macular optical coherence tomography (OCT) scans from 2 UK Biobank participants in the top 1% of the central serous chorioretinopathy genetic risk score (GRS) distribution. a) Broad irregular PED underneath the fovea (arrow) and b) RPE clumping (star).

### Effect Size Comparison Between CSC and AMD

We compared the allelic effect sizes of risk variants between CSC and AMD. At established AMD risk loci, allelic effects for AMD and CSC were inversely correlated (R = −0.78; P = 0.0043, Supplementary Figure S4). A reciprocal analysis of CSC loci showed a similar but nonsignificant trend (R = −0.41; P = 0.49). These results were consistent with a prior report (Rämö, 2023; de Jong et al., 2015)) 5/20/2026 1:09:00 PMwhich found that while CSC and AMD partially share susceptibility loci, the direction of effect is frequently opposite.

## Discussion

CSC remains an understudied cause of central vision loss and has an elusive pathophysiological basis despite its modest prevalence. The study of the anatomic origin of CSC has been hampered by the smaller size of prior genetic studies and the absence of systematic post-GWAS analyses capable of localizing disease risk to specific tissues and cell types. This study leverages 5 cohorts and 1,047,039 individuals to identify novel risk loci and is paired with an integrative post-GWAS framework. Our results converged on a model that CSC susceptibility is concentrated in scleral fibroblasts and vascular endothelial cells. While RPE abnormalities are certainly a clinical hallmark of CSC, these results support the classification of CSC as a disorder stemming first from the sclera and choroidal vasculature with RPE damage as a downstream consequence.

Our meta-analysis confirmed and expanded the catalog of CSC-associated loci. All previously reported associations in European cohorts were replicated. Notably, the loci harboring *GATA5* and *TNFRSF10A* that were previously identified in Asian cohorts were also replicated, confirming that at least a subset of CSC genetic architecture is shared across ancestries. Three novel genome-wide significant loci were identified, with nearest genes *TGFB1, LINC00551*, and *LOC105375630*. Between-study heterogeneity was low across all ten loci, supporting robust cross-cohort replicability and underscoring the likelihood of accurate phenotyping in these cohorts.

With integrative prioritization, we identified 10 putative causal genes (*CFH, CD46, AXL, PREX1, TGFB1, TNFRSF10A, PTPRB, GATA5, CASP10* and *NOTCH4*), each supported by at least 2 independent lines of genetic evidence (Figure 2c). Among these putative causal genes, MR and colocalization analyses pointed to 2 interesting candidates: *TGFB1* and *TNFRSF10A*. Colocalization demonstrated near-certain evidence of a shared causal variant between the TGFB1 pQTL and the CSC risk signal (PP4 = 0.999), supported by nominally significant MR estimates indicating that genetically predicted higher circulating TGFB1 levels are associated with lower CSC risk. TGFB1 is a master regulator of extracellular matrix deposition and fibroblast activation (Meng, Nikolic-Paterson and Lan, 2016), and pathological *TGFB1* signaling has been implicated in scleral thickness in mammalian myopia models (Jobling *et al*., 2004). Similarly, the TNFRSF10A pQTL and the CSC risk signal showed near-certain evidence of a shared causal variant (PP4 = 1.00), and MR implicated reduced circulating TNFRSF10A as a causal driver of CSC risk. TNFRSF10A encodes TRAIL receptor 1 which is expressed on endothelial cells and has established roles in regulating endothelial apoptosis and vascular permeability (Li et al., 2003; Secchiero et al., 2004). While further studies are needed to better articulate the role of TGFB1 and TNFRSF10A in CSC pathogenesis, circulating levels of both appear to be in the causal pathway to CSC and such levels are theoretically targetable.

In keeping with clinical observation, vascular endothelial cells were the predominant cellular carrier of CSC genetic risk across all 3 atlases. This endothelial signal is biologically coherent with several prioritized causal genes. *CFH* and *CD46*, both of which are regulators of the alternative complement pathway expressed on choroidal endothelial surfaces (Zipfel and Skerka, 2009; Kerr and Richards, 2012), represent the strongest overall associations in this study. Notably, while *CFH* variants are also among the strongest risk factors for AMD, the direction of effect is opposite across the 2 diseases, suggesting that CSC and AMD represent distinct pathophysiological responses to complement dysregulation. As noted, *TNFRSF10A* is expressed on endothelial cells and has established roles in regulating endothelial apoptosis and vascular permeability.

*PTPRB*, a receptor tyrosine phosphatase that regulates Tie-2 signaling and is critical for vascular integrity was again shown to harbor a rare variant associated with CSC, an association that was recently highlighted and shown to co-associate with varicose veins (Rämö, Gorman, *et al*., 2025b) and that prompted a clinical case series of faricimab in patients with CSC (Rämö, Kim, *et al*., 2025). It is important to note that the data here contain the data from the Rämo et al. single variant study (with the addition of the MGBB cohort and an updated AoU cohort). The association became stronger with the addition of these data.

The convergence of molecular and cellular evidence implicating sclera and vascular biology raises the question of whether such genetically defined risk is detectable as anatomical change on OCT. Stratifying UK Biobank participants by CSC polygenic risk and comparing macular OCT scans at the extremes of the risk distribution revealed a 2.7-fold enrichment of RPE abnormalities among genetically high-risk individuals. This indicates that aggregate genetic burden translates into detectable structural changes, suggesting that polygenic risk could complement imaging to help classify pachychoroid patients.

This study has limitations. First, our meta-analysis was composed of individuals of European ancestry, which may limit the generalizability of our findings to other populations with high CSC prevalence, particularly of Asian populations where prevalence remains high. Additionally, our enrichment analysis of cell types and anatomic regions are inherently subject to the dissection and labeling limitation of the cell atlases analyzed. The retinal and choroidal layers are both anatomically and functionally highly intertwined, and there are few atlases that have a high-resolution, layer-specific characterization of the posterior segment. This limitation motivated our inclusion of comprehensive ocular atlases such as Tabula Sapiens rather than restricting our examination to more specific posterior eye atlases. Moreover, our investigation of the individual contributions of arterial, venous, or choriocapillaris vasculature remains limited, as all endothelial cells were grouped together and lacked the dissection granularity to differentiate these anatomic distinctions.

In conclusion, this study provides an updated comprehensive genetic characterization of CSC and provides insights into disease pathogenesis. Continued work to understand the genetics of CSC subphenotypes, the interaction of genetics with known risk factors like steroid exposure and the role of yet un-analyzed variation including exome-sequencing data is ongoing.

## Data availability

The source data from FinnGen, UK Biobank and All of Us are available under restricted access due to the sensitive nature of the individual-level genotype and phenotype information. Individual-level genotypes and register data from FinnGen participants can be accessed by approved researchers via the Fingenious portal (https://site.ngenious./en/) hosted by the Finnish Biobank Cooperative FinBB (https://nbb./en/). Access to individual-level UKB data may be requested by researchers in academic, commercial, and charitable organizations (https://www.ukbiobank.ac.uk). This study used data from the All of Us Research Program’s Controlled Tier Dataset v8, available to authorized users on the Researcher Workbench (https://www.researchallofus.org). The raw Million Veteran Program data are protected and are not available due to data privacy laws. Summary-level association data from MVP used in this study are available through dbGaP, under accession code phs001672.v11.p1 [https://www.ncbi.nlm.nih.gov/projects/gap/cgi-bin/study.cgi?study_id=phs001672.v11.p1]. The meta-analysis summary statistics will be shared publicly after acceptance of this manuscript.

## Code availability

No custom software or analysis tools were created for this study; all analyses used existing software and tools as described in the Methods.

## Acknowledgements

We want to acknowledge the participants and investigators of the FinnGen study. We want to acknowledge the participants and investigators of the VA Million Veteran Program. Detailed acknowledgements for FinnGen and MVP are provided in Supplementary Information.

## Reference

1000 Genomes Project Consortium et al. (2015) “A global reference for human genetic variation,” Nature, 526(7571), pp. 68–74. Available at: 10.1038/nature15393.

All of Us Research Program Investigators et al. (2019) “The ‘All of Us’ Research Program,” The New England Journal of Medicine, 381(7), pp. 668–676. Available at: 10.1056/NEJMsr1809937.

Bergström, A. et al. (2020) “Insights into human genetic variation and population history from 929 diverse genomes,” Science, 367(6484), p. eaay5012. Available at: 10.1126/science.aay5012.

Brinks, J., van Dijk, E. H. C., Meijer, O. C., Schlingemann, R. O., & Boon, C. J. F. (2022). Choroidal arteriovenous anastomoses: A hypothesis for the pathogenesis of central serous chorioretinopathy and other pachychoroid disease spectrum abnormalities. Acta Ophthalmologica, 100(8), 946–959. 10.1111/aos.15112

Castro, V.M. et al. (2022) “The Mass General Brigham Biobank Portal: an i2b2-based data repository linking disparate and high-dimensional patient data to support multimodal analytics,” Journal of the American Medical Informatics Association: JAMIA, 29(4), pp. 643–651. Available at: 10.1093/jamia/ocab264.

Chang, C.C. et al. (2015) “Second-generation PLINK: rising to the challenge of larger and richer datasets,” GigaScience, 4, p. 7. Available at: 10.1186/s13742-015-0047-8.

Das, S. et al. (2016) “Next-generation genotype imputation service and methods,” Nature Genetics, 48(10), pp. 1284–1287. Available at: 10.1038/ng.3656.

van Dijk, E.H.C. et al. (2019) “FAMILIAL CENTRAL SEROUS CHORIORETINOPATHY,” Retina, 39(2), pp. 398–407. Available at: 10.1097/IAE.0000000000001966.

Fang, H. et al. (2019) “Harmonizing Genetic Ancestry and Self-identified Race/Ethnicity in Genome-wide Association Studies,” American Journal of Human Genetics, 105(4), pp. 763–772. Available at: 10.1016/j.ajhg.2019.08.012.

Feenstra, H. M. A., van Dijk, E. H. C., Cheung, C. M. G., Ohno-Matsui, K., Lai, T. Y. Y., Koizumi, H., Larsen, M., Querques, G., Downes, S. M., Yzer, S., Breazzano, M. P., Subhi, Y., Tadayoni, R., Priglinger, S. G., Pauleikhoff, L. J. B., Lange, C. A. K., Loewenstein, A., Diederen, R. M. H., Schlingemann, R. O., … Boon, C. J. F. (2024). Central serous chorioretinopathy: An evidence-based treatment guideline. Progress in Retinal and Eye Research, 101, 101236. 10.1016/j.preteyeres.2024.101236

Fritsche, L.G. et al. (2016) “A large genome-wide association study of age-related macular degeneration highlights contributions of rare and common variants,” Nature Genetics, 48(2), pp. 134–143. Available at: 10.1038/ng.3448.

Fung, A.T., Yang, Y. and Kam, A.W. (2023) “Central serous chorioretinopathy: A review,” Clinical & Experimental Ophthalmology, 51(3), pp. 243–270. Available at: 10.1111/ceo.14201.

Gaziano, J.M. et al. (2016) “Million Veteran Program: A mega-biobank to study genetic influences on health and disease,” Journal of Clinical Epidemiology, 70, pp. 214–223. Available at: 10.1016/j.jclinepi.2015.09.016.

Giambartolomei, C. et al. (2014) “Bayesian test for colocalisation between pairs of genetic association studies using summary statistics,” PLoS genetics, 10(5), p. e1004383. Available at: 10.1371/journal.pgen.1004383.

Hemani, G. et al. (2018) “The MR-Base platform supports systematic causal inference across the human phenome,” eLife, 7, p. e34408. Available at: 10.7554/eLife.34408.

Hosoda, Y. et al. (2019) “Genome-wide association analyses identify two susceptibility loci for pachychoroid disease central serous chorioretinopathy,” Communications Biology, 2, p. 468. Available at: 10.1038/s42003-019-0712-z.

Igo, R.P., Kinzy, T.G. and Cooke Bailey, J.N. (2019) “Genetic Risk Scores,” Current Protocols in Human Genetics, 104(1), p. e95. Available at: 10.1002/cphg.95.

Jobling, A.I. et al. (2004) “Isoform-specific changes in scleral transforming growth factor-beta expression and the regulation of collagen synthesis during myopia progression,” The Journal of Biological Chemistry, 279(18), pp. 18121–18126. Available at: 10.1074/jbc.M400381200.

de Jong, E. K., Breukink, M. B., Schellevis, R. L., Bakker, B., Mohr, J. K., Fauser, S., Keunen, J. E. E., Hoyng, C. B., den Hollander, A. I., & Boon, C. J. F. (2015). Chronic central serous chorioretinopathy is associated with genetic variants implicated in age-related macular degeneration. Ophthalmology, 122(3), 562–570. 10.1016/j.ophtha.2014.09.026

Kanda, P. et al. (2022) “Pathophysiology of central serous chorioretinopathy: a literature review with quality assessment,” Eye, 36(5), pp. 941–962. Available at: 10.1038/s41433-021-01808-3.

Kerr, H. and Richards, A. (2012) “Complement-mediated injury and protection of endothelium: lessons from atypical haemolytic uraemic syndrome,” Immunobiology, 217(2), pp. 195–203. Available at: 10.1016/j.imbio.2011.07.028.

Kitzmann, A.S. et al. (2008) “The incidence of central serous chorioretinopathy in Olmsted County, Minnesota, 1980-2002,” Ophthalmology, 115(1), pp. 169–173. Available at: 10.1016/j.ophtha.2007.02.032.

Koyama, S. et al. (2025) “Genetics and context for precision health in Greater Boston,” Nature Communications, 16(1), p. 11661. Available at: 10.1038/s41467-025-66598-8.

Kurki, M.I. et al. (2023) “FinnGen provides genetic insights from a well-phenotyped isolated population,” Nature, 613(7944), pp. 508–518. Available at: 10.1038/s41586-022-05473-8.

de Leeuw, C.A. et al. (2015) “MAGMA: generalized gene-set analysis of GWAS data,” PLoS computational biology, 11(4), p. e1004219. Available at: 10.1371/journal.pcbi.1004219.

Mägi, R. and Morris, A.P. (2010) “GWAMA: software for genome-wide association meta-analysis,” BMC bioinformatics, 11, p. 288. Available at: 10.1186/1471-2105-11-288.

Martin, F.J. et al. (2023) “Ensembl 2023,” Nucleic Acids Research, 51(D1), pp. D933–D941. Available at: 10.1093/nar/gkac958.

Mbatchou, J. et al. (2021) “Computationally efficient whole-genome regression for quantitative and binary traits,” Nature Genetics, 53(7), pp. 1097–1103. Available at: 10.1038/s41588-021-00870-7.

Meng, X.-M., Nikolic-Paterson, D.J. and Lan, H.Y. (2016) “TGF-β: the master regulator of fibrosis,” Nature Reviews. Nephrology, 12(6), pp. 325–338. Available at: 10.1038/nrneph.2016.48.

Menon, M. et al. (2019) “Single-cell transcriptomic atlas of the human retina identifies cell types associated with age-related macular degeneration,” Nature Communications, 10(1), p. 4902. Available at: 10.1038/s41467-019-12780-8.

Miki, A. et al. (2014) “Common variants in the complement factor H gene confer genetic susceptibility to central serous chorioretinopathy,” Ophthalmology, 121(5), pp. 1067–1072. Available at: 10.1016/j.ophtha.2013.11.020.

Monavarfeshani, A. et al. (2023) “Transcriptomic analysis of the ocular posterior segment completes a cell atlas of the human eye,” Proceedings of the National Academy of Sciences of the United States of America, 120(34), p. e2306153120. Available at: 10.1073/pnas.2306153120.

Mori, Y. et al. (2025) “Genome-wide association and multi-omics analyses provide insights into the disease mechanisms of central serous chorioretinopathy,” Scientific Reports, 15(1), p. 9158. Available at: 10.1038/s41598-025-92210-6.

Mulfaul, K. et al. (2022) “Local factor H production by human choroidal endothelial cells mitigates complement deposition: implications for macular degeneration,” The Journal of Pathology, 257(1), pp. 29–38. Available at: 10.1002/path.5867.

Pauleikhoff, L. J. B., Diederen, R. M. H., Chang-Wolf, J. M., Moll, A. C., Schlingemann, R. O., van Dijk, E. H. C., & Boon, C. J. F. (2024). Choroidal hyperpermeability patterns correlate with disease severity in central serous chorioretinopathy: CERTAIN study report 2. Acta Ophthalmologica, 102(6), e946–e955. 10.1111/aos.16679

Rämö, J.T. et al. (2023) “Overlap of Genetic Loci for Central Serous Chorioretinopathy With Age-Related Macular Degeneration,” JAMA ophthalmology, 141(5), pp. 449–457. Available at: 10.1001/jamaophthalmol.2023.0706.

Rämö, J.T., Gorman, B.R., et al. (2025) “Rare genetic variation in PTPRB is associated with central serous chorioretinopathy, varicose veins and glaucoma,” Nature Communications, 16(1), p. 4127. Available at: 10.1038/s41467-025-58686-6.

Rämö, J.T., Kim, L.A., et al. (2025) “Targeting the Tie-2 Receptor With Faricimab in Central Serous Chorioretinopathy: A Case Series Motivated by a Genetic Finding,” American Journal of Ophthalmology, 269, pp. 246–254. Available at: 10.1016/j.ajo.2024.08.040.

Schellevis, R.L. et al. (2018) “Role of the Complement System in Chronic Central Serous Chorioretinopathy: A Genome-Wide Association Study,” JAMA ophthalmology, 136(10), pp. 1128–1136. Available at: 10.1001/jamaophthalmol.2018.3190.

Sudlow, C. et al. (2015) “UK biobank: an open access resource for identifying the causes of a wide range of complex diseases of middle and old age,” PLoS medicine, 12(3), p. e1001779. Available at: 10.1371/journal.pmed.1001779.

Sun, B.B. et al. (2023) “Plasma proteomic associations with genetics and health in the UK Biobank,” Nature, 622(7982), pp. 329–338. Available at: 10.1038/s41586-023-06592-6.

Tabula Sapiens Consortium* et al. (2022) “The Tabula Sapiens: A multiple-organ, single-cell transcriptomic atlas of humans,” Science, 376(6594), p. eabl4896. Available at: 10.1126/science.abl4896.

Tokolyi, A. et al. (2025) “The contribution of genetic determinants of blood gene expression and splicing to molecular phenotypes and health outcomes,” Nature Genetics, 57(3), pp. 616–625. Available at: 10.1038/s41588-025-02096-3.

Wallace, C. (2020) “Eliciting priors and relaxing the single causal variant assumption in colocalisation analyses,” PLoS genetics, 16(4), p. e1008720. Available at: 10.1371/journal.pgen.1008720.

Wang, M. et al. (2008) “Central serous chorioretinopathy,” Acta Ophthalmologica, 86(2), pp. 126–145. Available at: 10.1111/j.1600-0420.2007.00889.x.

Weeks, E.M. et al. (2023) “Leveraging polygenic enrichments of gene features to predict genes underlying complex traits and diseases,” Nature Genetics, 55(8), pp. 1267–1276. Available at: 10.1038/s41588-023-01443-6.

Weenink, A.C., Borsje, R.A. and Oosterhuis, J.A. (2001) “Familial chronic central serous chorioretinopathy,” Ophthalmologica. Journal International D’ophtalmologie. International Journal of Ophthalmology. Zeitschrift Fur Augenheilkunde, 215(3), pp. 183–187. Available at: 10.1159/000050855.

Zhang, M.J. et al. (2022) “Polygenic enrichment distinguishes disease associations of individual cells in single-cell RNA-seq data,” Nature Genetics, 54(10), pp. 1572–1580. Available at: 10.1038/s41588-022-01167-z.

Zipfel, P.F. and Skerka, C. (2009) “Complement regulators and inhibitory proteins,” Nature Reviews. Immunology, 9(10), pp. 729–740. Available at: 10.1038/nri2620.

