## Supplemental Information for "GWAS Meta-analysis Identifies Novel Associated Loci and Points to Causal Tissues in Central Serous Chorioretinopathy"

**Supplementary Information**

**Study Cohorts**

The MGBB is a biorepository of over 200,000 consented patient samples at Mass General Brigham (parent organization of Massachusetts General Hospital and Brigham and Women’s Hospital, Boston, USA) (Koyama *et al.*, 2025). It was established in 2009 and enrollment is currently ongoing. In this study, we included 53,846 genotyped participants. Samples were genotyped using 3 versions of the biobank single nucleotide polymorphism (SNP) array offered by Illumina, which is designed to capture the diversity of genetic backgrounds across the globe. Details on arrays and quality control are provided in the Supplementary Methods. Samples were imputed to TOPMed r2 reference panel using the Michigan Imputation Server (Das *et al.*, 2016). Phenotypes were based on electronic health record data. EUR ancestry was assigned using a random forest classifier trained on principal components from the HGDP and 1000 Genomes reference panels, retaining individuals with a posterior probability > 0.5 for European ancestry.

FinnGen is a public-private partnership research project that combines genotype data from newly collected and legacy samples administered by Finnish biobanks (https://www.finngen.fi/en) to provide novel insight into human diseases. This study included genotype data from 486,484 individuals from FinnGen Data Freeze 12. The data were linked by unique national personal identification numbers to the national hospital discharge registry (available from 1968) and the specialist outpatient registry (1998–). EUR individuals were determined by first removing non-European outliers via principal component analysis (PCA) with 1000 Genomes, then retaining only individuals with ≥95% probability of belonging to the Finnish genetic cluster based on Mahalanobis distance.

The MVP is a population-scale biobank within the Department of Veterans Affairs. Voluntary enrollment of veterans receiving care in the Veterans Affairs (VA) began in 2011. All samples were scanned on the MVP 1.0 Axiom array (ThermoFisher). Samples were imputed to the TOPMed reference panel (version r2; N = 97,256 samples) using Minimac4 (v1.0.0). Phenotypes were based on electronic health record data with follow-up through the end of 2022. This study included genotype data from 273,904 individuals from MVP. Ancestries were inferred by integrating self-reported race/ethnicity with genetic PCA through the Harmonizing Genetic Ancestry and Self-identified Race/Ethnicity (HARE) algorithm's support vector machine (Fang *et al.*, 2019).

The AoU Research Program opened for enrollment in May 2018 and plans to enroll at least 1 million persons in the United States, collecting electronic health record data and biospecimens to advance the prevention and treatment of diseases. In the current project, we used genetic data from 228,707 short-read whole genome-sequencing samples in the v8 release and excluded participants without linked electronic health record data. AoU assigns ancestry using a Random Forest classifier trained on HGDP (Bergström *et al.*, 2020) and 1000 Genomes reference labels across 16 principal components (PCs), requiring ≥90% predicted probability for EUR classification.

The Dutch chronic-CSC (cCSC) cohort included 521 European patients recruited from the outpatient clinics of the Radboud University Medical Center (the Netherlands), University Hospital of Cologne (Germany), and Leiden University Medical Center (the Netherlands). Controls included 3,577 participants in the Nijmegen Biomedical Study. The patients with cCSC had subretinal fluid in at least one eye, RPE irregularities with characteristic leakage on fluorescein angiography and corresponding hyperfluorescence on indocyanine green angiography. Genotyping was performed with OmniExpress-12 or OmniExpress-24 chips, and data were imputed with the Haplotype Reference Consortium (release 1.1.2016).

UKB is a deeply phenotyped and genotyped prospective population-level cohort, which recruited approximately 500,000 participants aged 40–69 in the United Kingdom (UK) between 2006–2010 (Sudlow *et al.*, 2015). UKB participants were genotyped on the Applied Biosystems UK BiLEVE Axiom Array and the Applied Biosystems UKB Axiom Array (Affymetrix, US).

**Quality control of genomic data**

The All of Us short-read whole-genome sequencing data were pre-processed and quality-controlled centrally by the AoU Research Program, as described in the AoU Genomic Research Data Quality Report (https://support.researchallofus.org/hc/en-us/articles/29390274413716). Briefly, sample-level QC filtered on array–WGS fingerprint concordance (log-likelihood ratio > −3), sex concordance, cross-individual contamination rate (<3%), and sequencing coverage (mean coverage >30× with ≥90% of the genome covered at ≥20×), with additional removal of outliers based on per-sample variant metrics. Variant-level QC of the joint callset enforced genotype-level thresholds (genotype quality [GQ] ≥ 20, depth of coverage [DP] ≥ 10, and allele balance ≥ 0.2 at heterozygous calls), filters on excess heterozygosity (excessHet < 54.60), variant quality scores (QUAL < 60 for SNPs and < 69 for indels), excess alternate allele filters, and the AoU variant extract-train-score (VETS) filter; variants flagged by AoU were removed from downstream analyses. On top of the centralized AoU QC, we excluded any samples flagged as problematic by AoU and further filtered variants at a call rate <98%, Hardy–Weinberg equilibrium p-value in controls <1×10⁻⁸, and minor allele frequency (MAF) <1%. Principal components were computed within individuals of European ancestry using PLINK v1.9, and association testing was adjusted for sex, age, age², and the first ten principal components to account for residual population structure.

For the MGBB cohort QC, we excluded variants with minor allele frequency (MAF) <5%, missingness >5%, evidence of genotyping batch bias (p < 5×10⁻⁵), deviation from Hardy–Weinberg equilibrium at p < 1×10⁻¹⁰, and palindromic single nucleotide variants (A/T and C/G). At the sample level, we excluded individuals whose self-reported sex did not match their genetically inferred sex and individuals with an elevated heterozygosity ratio. Cleaned datasets were phased with SHAPEIT4 and imputed to the TOPMed r2 reference panel via the Michigan Imputation Server, and per-array imputed datasets were union-merged. Following imputation, we retained variants with imputation quality Rsq > 0.6 and post-imputation missingness < 5%. Principal components and continental genetic ancestry probabilities were derived by first intersecting common (MAF > 5%, genotyping rate > 95%), independent (R² < 0.1) variants between the HGDP/1000 Genomes reference panel and MGBB. PCs were calculated within HGDP/1000G samples and MGBB participants were projected into this PC space. A random forest classifier trained on the reference PCs and continental ancestry labels was then applied to MGBB, and individuals with a posterior probability > 0.5 for European ancestry were retained.

For the MVP cohort, genotyping array design, genotype calling, and the unified centralized quality-control pipeline applied to the MVP 1.0 Axiom array have been described in detail previously (Hunter-Zinck et al., 2020). Briefly, the unified QC pipeline applied to an initial release of 485,856 genotyped MVP participants retained 459,777 unique individuals and 668,418 high-quality markers, with high call performance across both common and rare variants.

The FinnGen genotype imputation protocol is available at: <https://dx.doi.org/10.17504/protocols.io.xbgfijw>. Estonian Biobank samples were genotyped and PLINK format files were created using Illumina GenomeStudio v2.0.4. Individuals were excluded from the analysis if their call-rate was < 95% or if sex defined based on heterozygosity of X chromosome did not match sex in phenotype data. Before imputation, variants were filtered by call-rate < 95%, HardyWeinberg Equilibrium p-value < 1e-4 (autosomal variants only), and minor allele frequency < 1%. Variant positions were in build 37 and all variants were changed to be from TOP strand using GSAMD-24v1-0_20011747_A1-b37.strand.RefAlt.zip files from https://www.well.ox.ac.uk/~wrayner/strand/ webpage. Pre-phasing was performed using the © 2023 American Medical Association. All rights reserved. Eagle v2.3 software. The number of conditioning haplotypes Eagle2 uses when phasing each sample was set to: --Kpbwt=20000 and imputation was performed using Beagle v.28Sep18.79 with effective population size ne=20,000.

Quality control of the Dutch cCSC cohort was performed in PLINK v1.90 separately within each batch and again after merging. Within each batch, samples were excluded if their genotyping call rate was <97% or if reported sex disagreed with genotype-inferred sex (71 controls and 7 cases removed), and variants were excluded if their call rate was <98%, if they deviated from Hardy–Weinberg equilibrium at p < 1×10⁻⁶, or if their minor allele frequency was <1%. The most recent case batch was lifted over from GRCh38 to GRCh37 to match the other batches using the UCSC LiftOver tool. Variants common to all batches were retained, the batches were merged, and strand inconsistencies were resolved by iterative flipping using the PLINK flipscan utility until concordance was reached across all datasets. Variants with a post-merge call rate <98% were then removed, leaving 589,487 autosomal and 13,282 X-chromosomal variants for downstream analysis. Population stratification was assessed by merging the cohort with HapMap3, pruning at a 50 kb window / step 5 / VIF 2, and principal-component analysis in PLINK; 2 controls and 13 cases identified as non-European outliers were excluded. Cryptic relatedness was evaluated with KING v2.0, and 4 controls and 5 cases exceeding a kinship coefficient of 0.0884 (duplicates and first- or second-degree relatives) were removed. After QC, 521 cCSC cases and 3,577 NBS controls remained. Autosomal variants were phased with Eagle v2.3 and X-chromosomal variants with SHAPEIT v2.r790, and the merged dataset was imputed to the Haplotype Reference Consortium reference panel (release 1.1.2016) using the Michigan Imputation Server. SNPs were filtered on an imputation quality score of R2 > 0.3 for common variants (MAF >5%) and a R2 > 0.8 for low frequency variants (MAF <5%).

**Gene-based association test**

We performed gene-based tests on the meta-analysis summary statistic Z-scores using MAGMA v1.10 (de Leeuw et al., 2015). SNPs were mapped to protein-coding genes following the Ensembl gene model (Martin et al., 2023). Gene-wise p-values were computed under the SNP-wise mean model using 1000 Genomes Phase 3 EUR as LD reference. MAF was set to a threshold of ≥0.001, and a 10kb window around gene start and stop. 6,535,749 SNPs (64.25%) were mapped to at least one gene, and SNPs were mapped to 17,832 genes. Gene-based p-values were calculated using the SNP-wise Mean model. Multiple testing was controlled by Bonferroni for gene-based tests (α = 0.05 / 17832).

**Gene prioritization with Polygenic Priority Score (PoPS)**

We prioritized putative effector genes using the PoPS (Weeks et al., 2023) framework applied to MAGMA gene-level association statistics. PoPS was run with the full feature panel distributed by the original PoPS study. Following the recommended PoPS workflow, we trained a ridge regression model with internal cross-validation on the genome-wide set of genes and generated out-of-fold PoPS scores for each gene to avoid overfitting. We retained only positively scored genes when ranking within loci. For locus-level gene nomination, we selected the top 2 PoPS-scoring genes within each locus to create a locus-specific candidate list.

**Colocalization analysis**

We performed Bayesian colocalization analysis using the coloc package (version 5.2.3) (Giambartolomei et al., 2014) to assess whether serum protein quantitative trait loci (pQTLs) from the UKB (Sun et al., 2023) and genetic variants associated with CSC risk share the same underlying causal variants. pQTL summary statistics were downloaded from Synapse (https://www.synapse.org/Synapse:syn51364943). We specified prior probabilities (p₁ = 1×10⁻⁴, p₂ = 1×10⁻⁴, and p₁₂ = 1×10⁻⁵) consistent with the typical genetic architecture of GWAS and molecular QTL studies (Wallace, 2020). The outputs of coloc are the probabilities of five mutually exclusive hypotheses for a genomic region (H₀: No association with either trait; H₁: Association with trait 1 only; H₂: Association with trait 2 only; H₃: Both traits associated, but with different causal variants; H₄: Both traits associated with the same shared causal variant). We defined significant colocalization as PP4/PP3 > 3 and PP4 ≥ 0.9. The PP4 threshold follows established practice (Wallace, 2020), while the additional PP4/PP3 ratio requirement reduces false-positive signals driven by residual uncertainty rather than clear differentiation from H₃ (Tokolyi et al., 2025).

**Mendelian Randomization (MR)**

We performed two-sample MR (Hemani et al., 2018a) to proteins with strong colocalization evidence, using pQTL from the UKB proteomics GWAS as exposures and our CSC meta-analysis as the outcome. For each protein, we restricted instruments to cis variants within the gene±100 kb window and required association evidence at p ≤ 0.01 in the pQTL summary statistics. To obtain approximately independent instruments, we LD-clumped the cis-eligible variants with PLINK 2.0 (Chang et al., 2015) against a 1000 Genomes European (hg38) reference panel (r² ≤ 0.10, 250 kb window, p1 = 5×10⁻⁸, p2 = 0.01). For MR estimation, we used inverse-variance weighted (IVW), simple median, and weighted median estimators. A protein was considered to be significant only when at least 2 of the 3 estimators yielded statistically significant estimates with concordant direction. We assessed directional horizontal pleiotropy using the MR-Egger intercept test. The MR-Egger slope was reported as a sensitivity estimate when ≥3 instruments are available. A non-significant MR-Egger intercept was interpreted as no evidence of directional horizontal pleiotropy. Multiple testing was controlled by Bonferroni (α < 0.05) across the number of significantly colocalized proteins.

**Cell type enrichment across single-cell RNA-seq datasets (scDRS)**

We assessed cellular enrichment of CSC genetic signals using the Single-Cell Disease Relevance Score (scDRS) method (Zhang et al., 2022). The prioritized set of genes from PoPS, colocalization and MR were used as input. Three independent human datasets were used: a retinal atlas (Menon et al., 2019), an ocular posterior segment atlas (Monavarfeshani et al., 2023), and the multi-organ Tabula Sapiens resource (Tabula Sapiens Consortium* et al., 2022).

For scDRS scoring, we used the full score mode with b = 5,000 matched control gene sets per trait, mean–variance control matching (--ctrl-match-opt mean_var), and variance-stabilized weighting (--weight-opt vs). Covariates included donor identifier and sex for all datasets; where available we additionally regressed out technical covariates (e.g., number of detected genes, total counts, and mitochondrial percentage). We required a minimum of 50 cells per group for inference. The primary inferential endpoint was the group-level association test (“assoc”) implemented in scDRS. Multiple testing was controlled using Benjamini–Hochberg FDR within each dataset and grouping variable (e.g., within cell-type labels or tissue compartments).

**FinnGen Ethics statement and materials & methods**

Study subjects in FinnGen provided informed consent for biobank research, based on the Finnish Biobank Act. Alternatively, separate research cohorts, collected prior the Finnish Biobank Act came into effect (in September 2013) and start of FinnGen (August 2017), were collected based on study-specific consents and later transferred to the Finnish biobanks after approval by Fimea (Finnish Medicines Agency), the National Supervisory Authority for Welfare and Health. Recruitment protocols followed the biobank protocols approved by Fimea. The Coordinating Ethics Committee of the Hospital District of Helsinki and Uusimaa (HUS) statement number for the FinnGen study is Nr HUS/990/2017.

The FinnGen study is approved by Finnish Institute for Health and Welfare (permit numbers: THL/2031/6.02.00/2017, THL/1101/5.05.00/2017, THL/341/6.02.00/2018, THL/2222/6.02.00/2018, THL/283/6.02.00/2019, THL/1721/5.05.00/2019 and THL/1524/5.05.00/2020), Digital and population data service agency (permit numbers: VRK43431/2017-3, VRK/6909/2018-3, VRK/4415/2019-3), the Social Insurance Institution (permit numbers: KELA 58/522/2017, KELA 131/522/2018, KELA 70/522/2019, KELA 98/522/2019, KELA 134/522/2019, KELA 138/522/2019, KELA 2/522/2020, KELA 16/522/2020), Findata permit numbers THL/2364/14.02/2020, THL/4055/14.06.00/2020, THL/3433/14.06.00/2020, THL/4432/14.06/2020, THL/5189/14.06/2020, THL/5894/14.06.00/2020, THL/6619/14.06.00/2020, THL/209/14.06.00/2021, THL/688/14.06.00/2021, THL/1284/14.06.00/2021, THL/1965/14.06.00/2021, THL/5546/14.02.00/2020, THL/2658/14.06.00/2021, THL/4235/14.06.00/2021, Statistics Finland (permit numbers: TK-53-1041-17 and TK/143/07.03.00/2020 (earlier TK-53-90-20) TK/1735/07.03.00/2021, TK/3112/07.03.00/2021) and Finnish Registry for Kidney Diseases permission/extract from the meeting minutes on 4^th^ July 2019.

The Biobank Access Decisions for FinnGen samples and data utilized in FinnGen Data Freeze 12 include: THL Biobank BB2017_55, BB2017_111, BB2018_19, BB_2018_34, BB_2018_67, BB2018_71, BB2019_7, BB2019_8, BB2019_26, BB2020_1, BB2021_65, Finnish Red Cross Blood Service Biobank 7.12.2017, Helsinki Biobank HUS/359/2017, HUS/248/2020, HUS/430/2021 §28, §29, HUS/150/2022 §12, §13, §14, §15, §16, §17, §18, §23, §58, §59, HUS/128/2023 §18, Auria Biobank AB17-5154 and amendment #1 (August 17 2020) and amendments BB_2021-0140, BB_2021-0156 (August 26 2021, Feb 2 2022), BB_2021-0169, BB_2021-0179, BB_2021-0161, AB20-5926 and amendment #1 (April 23 2020) and it´s modifications (Sep 22 2021), BB_2022-0262, BB_2022-0256, Biobank Borealis of Northern Finland_2017_1013, 2021_5010, 2021_5010 Amendment, 2021_5018, 2021_5018 Amendment, 2021_5015, 2021_5015 Amendment, 2021_5015 Amendment_2, 2021_5023, 2021_5023 Amendment, 2021_5023 Amendment_2, 2021_5017, 2021_5017 Amendment, 2022_6001, 2022_6001 Amendment, 2022_6006 Amendment, 2022_6006 Amendment, 2022_6006 Amendment_2, BB22-0067, 2022_0262, 2022_0262 Amendment, Biobank of Eastern Finland 1186/2018 and amendment 22§/2020, 53§/2021, 13§/2022, 14§/2022, 15§/2022, 27§/2022, 28§/2022, 29§/2022, 33§/2022, 35§/2022, 36§/2022, 37§/2022, 39§/2022, 7§/2023, 32§/2023, 33§/2023, 34§/2023, 35§/2023, 36§/2023, 37§/2023, 38§/2023, 39§/2023, 40§/2023, 41§/2023, Finnish Clinical Biobank Tampere MH0004 and amendments (21.02.2020 & 06.10.2020), BB2021-0140 8§/2021, 9§/2021, §9/2022, §10/2022, §12/2022, 13§/2022, §20/2022, §21/2022, §22/2022, §23/2022, 28§/2022, 29§/2022, 30§/2022, 31§/2022, 32§/2022, 38§/2022, 40§/2022, 42§/2022, 1§/2023, Central Finland Biobank 1-2017, BB_2021-0161, BB_2021-0169, BB_2021-0179, BB_2021-0170, BB_2022-0256, BB_2022-0262, BB22-0067, Decision allowing to continue data processing until 31^st^ Aug 2024 for projects: BB_2021-0179, BB22-0067,BB_2022-0262, BB_2021-0170, BB_2021-0164, BB_2021-0161, and BB_2021-0169, and Terveystalo Biobank STB 2018001 and amendment 25^th^ Aug 2020, Finnish Hematological Registry and Clinical Biobank decision 18^th^ June 2021, Arctic biobank P0844: ARC_2021_1001.

**Phenotype ascertainment**

In the MGBB cohort, patients with CSC were initially identified based on at least one instance of the ICD-10-CM code H35.71* or ICD-9-CM code 362.41. CSC diagnoses were subsequently validated through manual review of OCTs (and fluorescein angiography or indocyanine green angiography when available) by a retina specialist, and only those consistent with CSC were used as cases. Participants with AMD (ICD-10: H35.1, H35.2*, H35.3*; ICD-9: 362.5, 362.51, 362.52) were excluded.

In the All of Us cohort, patients with CSC were identified based on at least one instance of the ICD-10-CM code H35.71* or ICD−9-CM code 362.41, and all participants with age-related macular degeneration (AMD,ICD-10: H35.1*, H35.2*, H35.3*; ICD-9: 362.5, 362.51, 362.52) were excluded.

Case definitions for FinnGen, MVP, and the European chronic CSC cohort have been described previously (Rämö, Gorman, et al., 2025) and are summarized in Supplementary Data 1. Participant demographics, sex distribution, age at end of follow-up, and prevalence of pre-specified ocular comorbidities in the two newly analyzed biobank cohorts are provided in Supplementary Data 2.

**Acknowledgements of FinnGen study**

The FinnGen project is funded by two grants from Business Finland (HUS 4685/31/2016 and UH 4386/31/2016) and the following industry partners: AbbVie Inc., Alnylam Pharmaceuticals, Inc., AstraZeneca UK Ltd, Bayer AG, Biogen MA Inc., Boehringer Ingelheim International GmbH, Bristol Myers Squibb Inc. (and Celgene Corporation & Celgene International II Sàrl), Genentech Inc., GlaxoSmithKline Intellectual Property Development Ltd., Johnson&Johnson Innovative Medicine Inc., Maze Therapeutics Inc., Merck Sharp & Dohme LCC, Novartis AG, Pfizer Inc. and Sanofi US Services Inc. Following biobanks are acknowledged for delivering biobank samples to FinnGen: Auria Biobank (www.auria.fi/biopankki), THL Biobank (www.thl.fi/biobank), Helsinki Biobank (www.helsinginbiopankki.fi), Biobank Borealis of Northern Finland (https://www.ppshp.fi/Tutkimus-ja-opetus/Biopankki/Pages/Biobank-Borealis-briefly-in-English.aspx), Finnish Clinical Biobank Tampere (www.tays.fi/en-US/Research_and_development/Finnish_Clinical_Biobank_Tampere), Biobank of Eastern Finland (www.ita-suomenbiopankki.fi/en), Central Finland Biobank (www.ksshp.fi/fi-FI/Potilaalle/Biopankki), Finnish Red Cross Blood Service Biobank (www.veripalvelu.fi/verenluovutus/biopankkitoiminta), Terveystalo Biobank (www.terveystalo.com/fi/Yritystietoa/Terveystalo-Biopankki/Biopankki/) and Arctic Biobank (https://www.oulu.fi/en/university/faculties-and-units/faculty-medicine/northern-finland-birth-cohorts-and-arctic-biobank). All Finnish Biobanks are members of BBMRI.fi infrastructure (https://www.bbmri-eric.eu/national-nodes/finland/). Finnish Biobank Cooperative -FINBB (https://finbb.fi/) is the coordinator of BBMRI-ERIC operations in Finland. The Finnish biobank data can be accessed through the Fingenious® services (https://site.fingenious.fi/en/) managed by FINBB.

**Core acknowledgements for VA Million Veteran Program**

MVP Program Office

- Sumitra Muralidhar, Ph.D., Program Director

US Department of Veterans Affairs, 810 Vermont Avenue NW, Washington, DC 20420

- Jennifer Moser, Ph.D., Associate Director, Scientific Programs

US Department of Veterans Affairs, 810 Vermont Avenue NW, Washington, DC 20420

- Jennifer E. Deen, B.S., Associate Director, Cohort & Public Relations

US Department of Veterans Affairs, 810 Vermont Avenue NW, Washington, DC 20420

MVP Steering Committee

- Co-Chair: Philip S. Tsao, Ph.D.

VA Palo Alto Health Care System, 3801 Miranda Avenue, Palo Alto, CA 94304

- Co-Chair: Sumitra Muralidhar, Ph.D.

US Department of Veterans Affairs, 810 Vermont Avenue NW, Washington, DC 20420

- J. Michael Gaziano, M.D., M.P.H.

VA Boston Healthcare System, 150 S. Huntington Avenue, Boston, MA 02130

- Adriana Hung, M.D., M.P.H.,

VA Tennessee Valley Healthcare System, 1310 24th Avenue, South Nashville, TN 37212

- Dave Oslin, M.D.

Philadelphia VA Medical Center, 3900 Woodland Avenue, Philadelphia, PA 19104

- Deepak Voora, M.D.

Durham VA Medical Center, 508 Fulton Street, Durham, NC 27705

MVP Co-Principal Investigators

- J. Michael Gaziano, M.D., M.P.H.

VA Boston Healthcare System, 150 S. Huntington Avenue, Boston, MA 02130

- Philip S. Tsao, Ph.D.

VA Palo Alto Health Care System, 3801 Miranda Avenue, Palo Alto, CA 94304

MVP Core Operations

- Jessica V. Brewer, M.P.H., Director, MVP Cohort Operations

VA Boston Healthcare System, 150 S. Huntington Avenue, Boston, MA 02130

- Mary T. Brophy M.D., M.P.H., Director, VA Central Biorepository

VA Boston Healthcare System, 150 S. Huntington Avenue, Boston, MA 02130

- Kelly Cho, M.P.H, Ph.D., Director, MVP Phenomics

VA Boston Healthcare System, 150 S. Huntington Avenue, Boston, MA 02130

- Lori Churby, B.S., Director, MVP Regulatory Affairs

VA Palo Alto Health Care System, 3801 Miranda Avenue, Palo Alto, CA 94304

- Jacob T. Kean, Ph.D., Acting Director, VA Informatics and Computing Infrastructure (VINCI)

VA Salt Lake City Health Care System, 500 Foothill Drive, Salt Lake City, UT 84148

- Saiju Pyarajan Ph.D., Director, Data and Computational Sciences

VA Boston Healthcare System, 150 S. Huntington Avenue, Boston, MA 02130

- Robert Ringer, Pharm.D., Director, VA Albuquerque Central Biorepository

New Mexico VA Health Care System, 1501 San Pedro Drive SE, Albuquerque, NM 87108

- Luis E. Selva, Ph.D., Director, MVP Biorepository Coordination

VA Boston Healthcare System, 150 S. Huntington Avenue, Boston, MA 02130

- Shahpoor (Alex) Shayan, M.S., Director, MVP PRE Informatics

VA Boston Healthcare System, 150 S. Huntington Avenue, Boston, MA 02130

- Brady Stephens, M.S., Principal Investigator, MVP Information Center

Canandaigua VA Medical Center, 400 Fort Hill Avenue, Canandaigua, NY 14424

- Stacey B. Whitbourne, Ph.D., Director, MVP Cohort Development and Management

VA Boston Healthcare System, 150 S. Huntington Avenue, Boston, MA 02130
