## Supplemental Figures for "GWAS Meta-analysis Identifies Novel Associated Loci and Points to Causal Tissues in Central Serous Chorioretinopathy"

**Supplementary Figures**

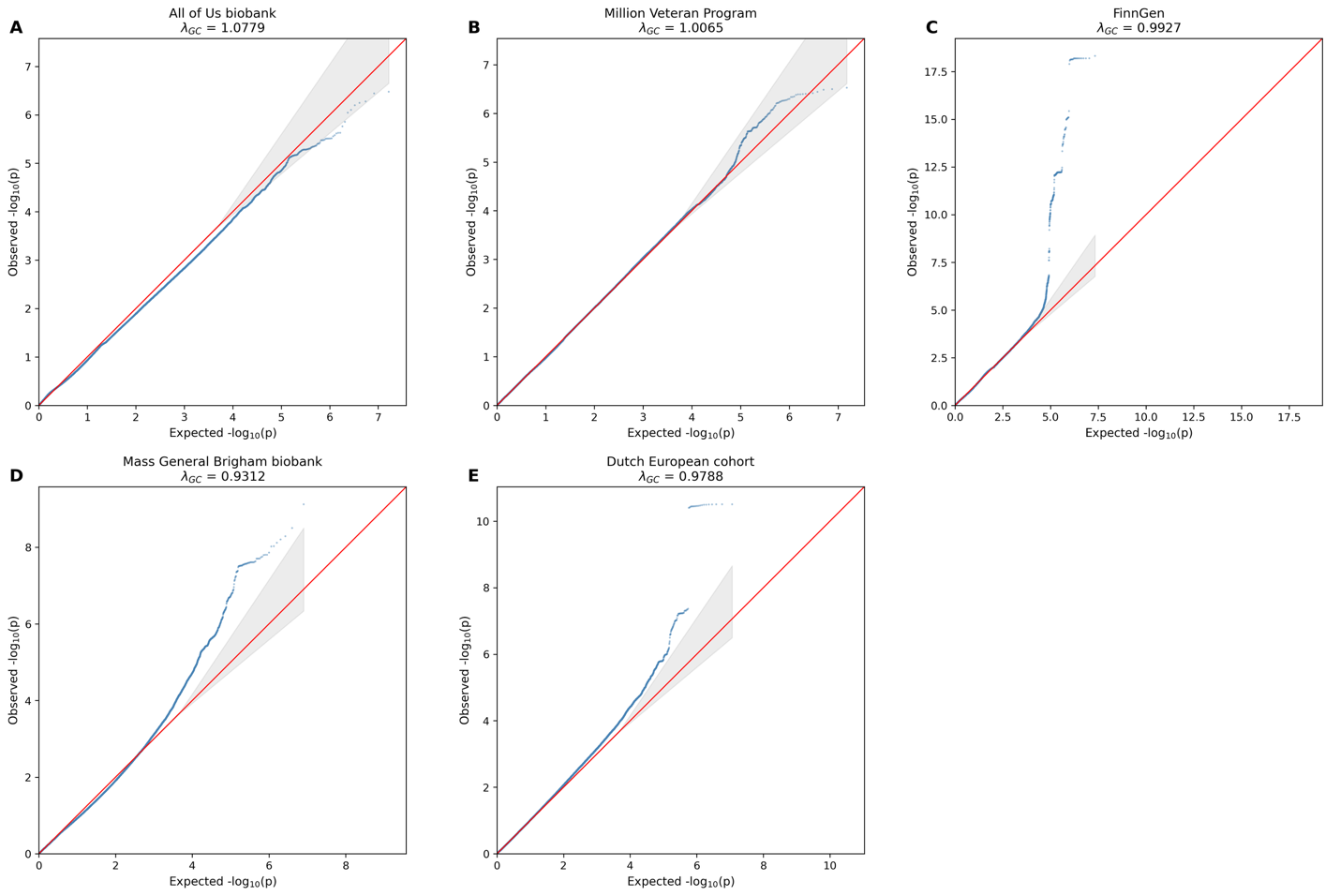

**Supplementary Figure 1.** **Quantile-Quantile (Q-Q) Plots of Individual Cohort GWAS Results.** Plots display the observed versus expected -log{10}(p) values for the five independent cohorts contributed to the meta-analysis: A, All of Us biobank; B, Million Veteran Program; C, FinnGen; D, Mass General Brigham Biobank; and E, the Dutch European cohort. The genomic inflation factor (lambda_{GC}) is provided for each cohort. The red line indicates the null hypothesis of no association (slope = 1), and the shaded gray region represents the 95% confidence interval. The lambda_{GC} values near 1.0 indicate adequate control for population stratification and cryptic relatedness prior to meta-analysis.

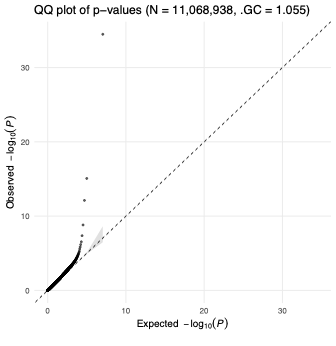

**Supplementary Figure S2.** Quantile-Quantile (Q-Q) plot of the CSC meta-analysis.

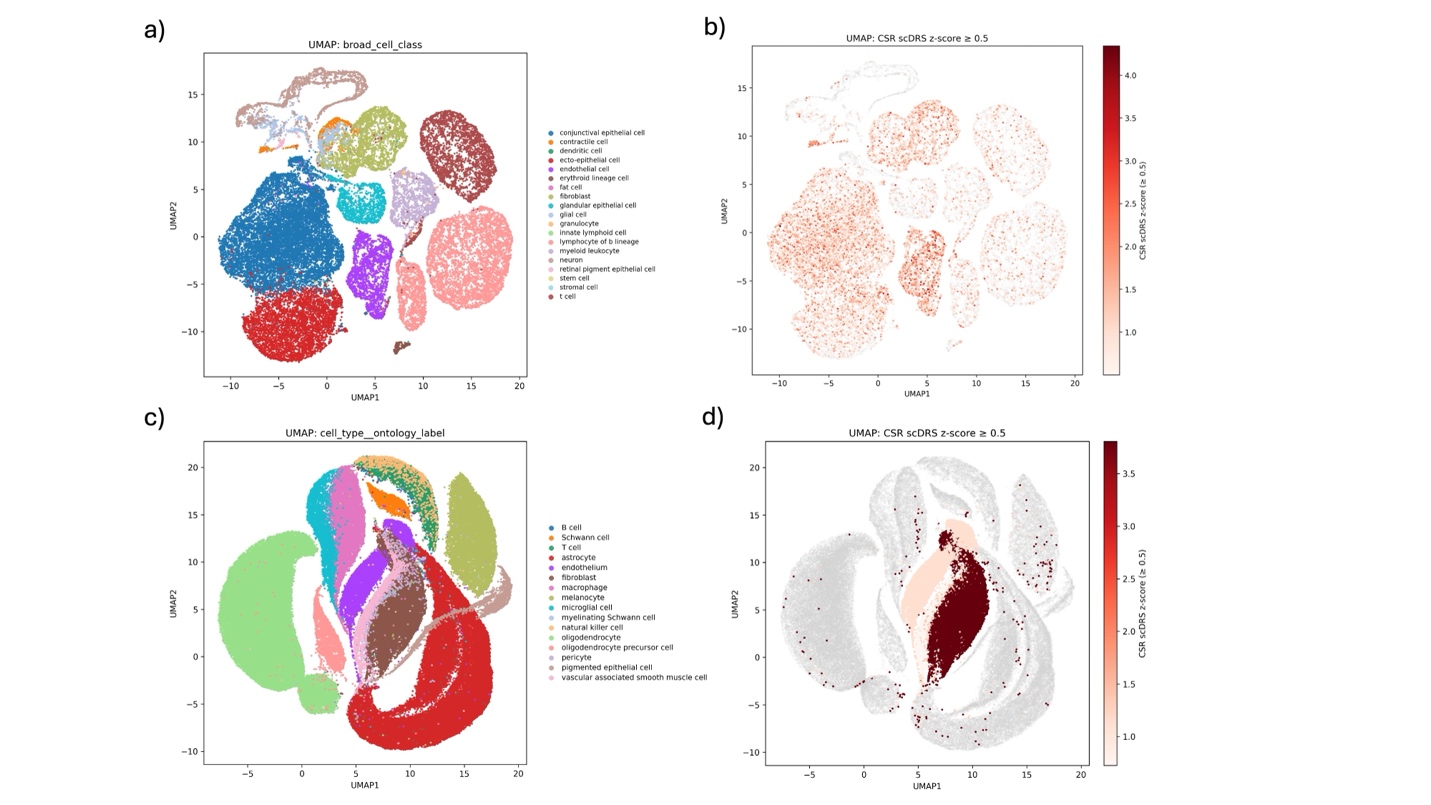

**Supplementary Figure S3. Uniform Manifold Approximation and Projection (UMAP) plots visualizing the enrichment of CSC genetic risk within specific cell populations.** a) UMAP of the Tabula Sapiens multi-organ atlas (ocular subset) colored by annotated cell type. b) The same UMAP colored by CSC scDRS z-score. Darker red indicates a higher aggregate genetic risk score for that individual cell. Note the elevated signal in endothelial and fibroblast clusters. c) UMAP of the Ocular Posterior Segment Atlas (Monavarfeshani et al.) colored by annotated cell type. d) The same UMAP colored by CSC scDRS z-score. The strongest signal (dark red) corresponds to the endothelial cell cluster (purple in panel C), indicating that CSC risk variants are preferentially active in vascular endothelium.

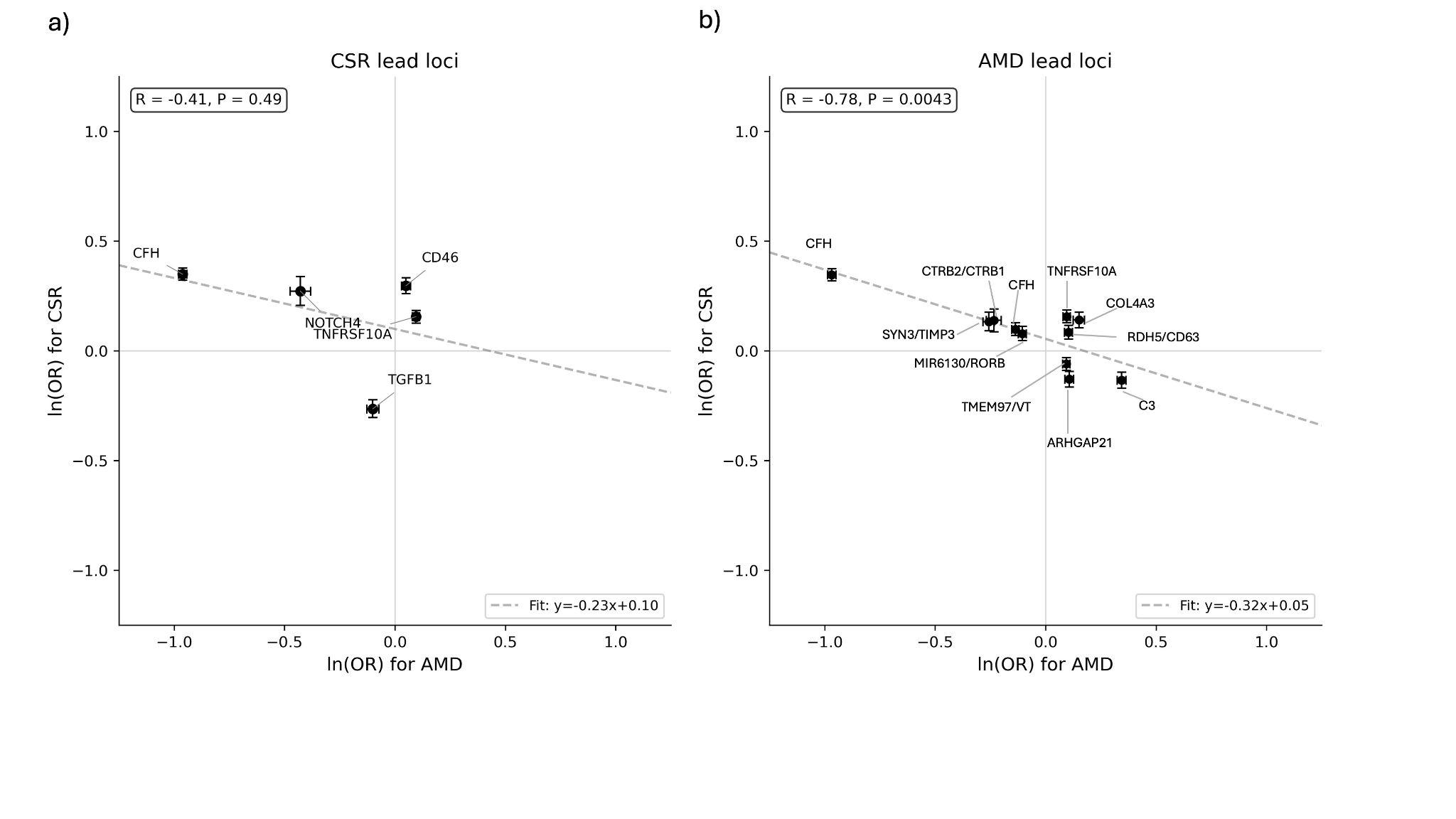

**Supplementary Figure S4. Scatterplots comparing the log-odds ratios (Beta) of risk variants for CSC (y-axis) against AMD (x-axis).** a) Correlation of effect sizes at the genome-wide significant CSC risk loci identified in this study. A similar negative trend is observed (R = -0.41), though it does not reach statistical significance (P = 0.49) likely due to the smaller number of available index variants. Error bars represent standard errors. b) Correlation of effect sizes at established AMD risk loci (Fritsche et al.). A significant negative correlation (R = -0.78; P = 0.0043) indicates that variants increasing risk for AMD are generally protective for CSC.
